# Intracranial Aneurysm Wall Displacement Predicts Instability

**DOI:** 10.1101/2022.06.02.22275917

**Authors:** A. Pionteck, J. Abderezaei, P. Fillingham, Y.-C. Chuang, Y. Sakai, P. Belani, B. Rigney, R. De Leacy, J. Fifi, A. Chien, P. Villablanca, G. Colby, R. Jahan, G. Duckwiler, J. Sayre, S. Holdsworth, M. Levitt, J. Mocco, M. Kurt, K. Nael

**Affiliations:** Department of Mechanical Engineering, University of Washington, Seattle, USA; Icahn School of Medicine at Mount Sinai, New York, USA; Diagnostic, Molecular and Interventional Radiology, Icahn School of Medicine at Mount Sinai, New York, USA; Department of Neurosurgery, Icahn School of Medicine at Mount Sinai, New York, USA; Department of Neurology, Icahn School of Medicine at Mount Sinai, New York, USA; Department of Radiology, University of California, Los Angeles, USA; Department of Neurosurgery, University of California, Los Angeles, USA; Department of Anatomy and Medical Imaging Centre for Brain Research, Faculty of Medical and Health Sciences, University of Auckland, Auckland, New Zealand; Medical Research Institute, Gisborne-Tairāwhiti, New Zealand; Departments of Neurological Surgery, Radiology, Mechanical Engineering, and Stroke & Applied Neuroscience Center, University of Washington School of Medicine, Seattle, USA

**Author notes:** These authors contributed equally to this work (A. Pionteck, J. Abderezaei).

**Keywords:** Intracranial Aneurysm, Instability Risk, Wall Deformation, aFlow, amplification, MRI

## Abstract

Ruptured intracranial aneurysms (IAs) are catastrophic events associated with a high mortality rate. An estimation of 6 million people in the United States have reported IAs, raising a pressing need for diagnostic tools to assess IAs rupture risks. Current population-based guidelines are imperfect, hence the need for new quantifiable variables and imaging markers. Aneurysm wall motion has been identified as a potential marker of high risk aneurysms, but conventional imaging techniques are challenged by small IAs sizes and limited spatial resolution. Recently, amplified Flow (aFlow) has been introduced as an algorithm which allows visualization and quantification of aneurysm wall motion based on amplification of 4D flow MRI data. In this work, we used aFlow to assess IAs wall motion in patients with growing aneurysms. The results were compared with a patient cohort with stable aneurysms. Among 118 patients with unruptured IAs who underwent sequential surveillance imaging, 10 patients with growing IAs who had baseline 3D TOF-MRA and 4D flow MR imaging were identified and matched with another cohort of patients with stable IAs based on IAs size and location. aFlow was then applied to the 4D flow MR data to amplify the aneurysm wall displacement. Voxel-based values of displacement were extracted for each aneurysm and normalized with respect to the reference parent artery. Following histogram analysis, the highest and lowest IAs displacements were calculated, together with their standard deviation and interquartile ranges. A paired-wise analysis was adopted to assess the differences among clinical variables, demographic data, morphological features, and aFlow parameters between patients with stable versus growing aneurysm. Results demonstrated higher wall motion and higher variability of deformation for the growing aneurysms, possibly due to inhomogeneities of the mechanical characteristics of the vessels walls or to underlying hemodynamics. Computational Fluid Dynamic simulation was also conducted for a subset of 6 stable and 6 growing aneurysms to examine the correlation between hemodynamic parameters, wall motion, and aneurysm stability. The magnitude and variance of directional wall shear stress gradient, in addition to area of colocation of elevated oscillatory shear stress and high variance in pressure, were highly correlated with both wall motion and aneurysm stability. We demonstrated here that the measurement and amplification of the aneurysm wall motion achieved with our method has the potential to differentiate stable from growing aneurysms, and potentially act as a substitute for in depth computational fluid dynamic analysis.

## 1. Introduction

There is a pressing need for a diagnostic tool able to assess the risk intracranial aneurysms (IAs) rupture. The reported incidence of IAs is approximately 2-5% in the general population with an estimation of 6 million people in the United States [1–4]. Even if the overall rupture rate for IAs is relatively small, ranging from 0.1% to 1.4% per year depending on associated risk factors [5–7], ruptured IAs are catastrophic events with a mortality rate ranging from 32-67% and significant morbidity in one third of the survivors [3, 8, 9].

Due to low rate of rupture, preventive treatment including surgical clipping and endovascular coiling is often considered in the context of the natural history of IAs with a strategy to carefully balance the risk of rupture against the risk associated with treatment complications and cost [4, 6, 10, 11]. Risk factors associated with growing aneurysms have been studied extensively; some of these include age, sex, family history of aneurysm or subarachnoid hemorrhage, smoking, hypertension, aneurysm size and location [6, 12– 15]. These population-based data have been used to create management and treatment guidelines that are used for risk stratification and treatment decision making such as PHASES score (population, hypertension, age, size of aneurysm, earlier subarachnoid hemorrhage from another aneurysm, site of aneurysm) [6, 16].

However, these population-based guidelines are rather imperfect in determining risk of rupture. For example IAs size of greater than 7 mm has been used as an objective measure of risk stratification with mixed results since a significant number of ruptured IAs can be small [17–19].

Therefore, other quantifiable variables and imaging markers have been investigated for more specific and individualized IA risk stratification such as presence of IA wall inflammation assessed by MR-vessel wall imaging [20, 21] and patient-specific hemodynamic variables obtained through computational fluid dynamic (CFD) studies [22–25]. Computational fluid dynamics shows great promise for quantifying hemodynamic parameters that are associated with aneurysm growth and rupture. Hemodynamic parameters that can only be captured through CFD, such as Oscillatory Shear Index and Wall Shear Stress Gradient, have been established as indicators for aneurysm development [26] and rupture risk [27] While CFD has the potential to provide invaluable insight to the study of aneurysm development and rupture, its clinical application is currently limited by the high level of engineering expertise necessary, a lack of methodological consistency between studies, and difficulty of obtaining large sample sizes for longitudinal study [28].

Aneurysm wall motion is shown as a potential marker to identify high risk aneurysms, where inhomogeneous mechanical characteristics and wall motion abnormalities are often associated with IAs growth and rupture [29–33]. Subjective parameters of irregular wall motion and deformation such as irregular pulsation and pulsating blebs were reported in growing or ruptured IAs [31, 32]. More objective and quantitative approaches in estimating IAs wall motion using dynamic cardiac gated magnetic resonance angiography (MRA) [34, 35] or computed tomography angiography (CTA) [36, 37] have been explored in determination of high risk IAs with some success. However, aneurysm wall motion has proven difficult to quantify with conventional imaging techniques due to the small size of IAs and limited spatial resolution. Small IAs wall deformations are often close to the imaging resolution, rendering quantitative wall motion analysis challenging [33].

Recently, a promising image processing algorithm called amplified Flow (aFlow) has been introduced which allows for visualization and quantification of aneurysm wall motion [38]. This amplification algorithm derives from an improved version of a Eulerian video magnification (EVM) algorithm [39–41]. By using a modal decomposition technique called dynamic mode decomposition (DMD) to capture the transient phenomena in the desired frequency range, arterial wall motion in a 4D flow MRI acquisition can be amplified and quantified [38]. In this study, we aimed to use the aFlow algorithm to assess IAs wall motion in a group of patients with growing aneurysms and perform a comparative analysis to a matched cohort with stable aneurysms. We then conducted CFD simulation on a sub cohort of these growing and stable aneurysms, allowing for direct correlation of wall motion and computationally derived hemodynamic parameters.

## 2. Methods

### 2.1. Human subjects

This retrospective case control study was approved by our institutional review board. Study subjects were identified within a prospectively collected data of patients with unruptured IAs who underwent MRA and 4D flow MRI between Jan 2016-Dec 2020. As part of this study, patients agreed to come back for follow up visits and undergo follow up MRA scans to determine interval change in aneurysm size. Informed consent was obtained from all individuals. The inclusion criteria were 1) Availability of MRA and 4D flow MRI at baseline imaging 2) Follow up MRA at least one year or later from the baseline imaging. At the time of analysis for this project, a total of 118 patients have been consented and enrolled who met the inclusion criteria.

### 2.2. Aneurysm measurement

IA size was measured independently by two board certified neuroradiologists with 10 and 6 years of experience. 3D time-of-flight MRA (TOF-MRA) source data were available in a commercially available software (Vitrea, Vital Images) and 3D multi-planar reformations were used to calculate the following in each IA: 1) Aneurysm size: maximum perpendicular distance of the dome from the neck plane; 2) Aneurysm neck: maximum diameter of the aneurysm neck where it attached to the parent vessel; 3) Aspect ratio: aneurysm size/neck. Individual measurements were first obtained on the baseline scan, then on the follow-up MRAs of the same patient. The observers were not blinded to the time order of the scans and had the baseline for comparison to mimic standard clinical practice. Among 118 patients enrolled, a total of 14 patients were identified as having growing aneurysms determined by follow up MRA studies. Determination of growth was based on interval increase in aneurysm size by 1mm [42] on two sequential follow up MRA. In four patients, the 4D flow MRI datasets were non diagnostic (motion, n=2; failed flow-encoding, n=2) resulting in a total of 10 patients with growing IAs for analysis. Subsequently a cohort of 10 patients with IAs whom aneurysm remained stable in size throughout the course of the study were selected and matched on the basis of aneurysm size and location. These twenty patients were included in the study for a case control pairwise analysis.

### 2.3. Imaging protocols

Image acquisition was performed on a 3T MR750 Discovery scanner (GE Healthcare, Milwaukee, WI, USA) or a 3T Skyra scanner (Siemens Healthineers AG, Germany). A multichannel head-neck coil with 20 elements were available on both scanners for signal reception. A TOF-MRA was obtained with identical sequence parameters on both scanners with the following parameters: TR/TE: 16/3.1 ms, FA: 20°, slices: 40, slice thickness: 0.6 mm, 4-axial slabs, matrix 384 × 384, FOV: 180 × 180 mm. For 4D flow MRI, a 3D phase-contrast peripherally-gated sequence was used with the following parameters: TR/TE = 5.8/3.1 ms, FA: 14°, matrix: 224 × 224, FOV: 180 mm, slices: 60-90, slice thickness: 1 mm. Twenty cardiac phases were obtained with a temporal resolution of approximately 40-60 ms (depending on the subject’s heart rate). A 3D-velocity encoding value (venc) of 80 cm/s was chosen [43]. Sequence parameters between GE and Siemens scanners varied slightly due to technical differences and restrictions imposed by the scanner hardware, however the voxel-sizes were kept near identical throughout the study.

### 2.4. Preprocessing of the 4D flow MRI data

The acquired images underwent a series of preprocessing steps to improve their quality before amplification (Figure 1). The raw data was corrected for background phase errors arising from Maxwell terms [44, 45], gradient field non-linearity [45, 46], and eddy currents [47]. Phase errors due to eddy currents were corrected by fitting a third-order polynomial to the static regions of the image and subsequently subtracting this from the velocity data [47]. Finally, to visualize the blood vessels, the acquired anatomical magnitude data was combined with the 3D velocity information [48].

**Figure 1:**
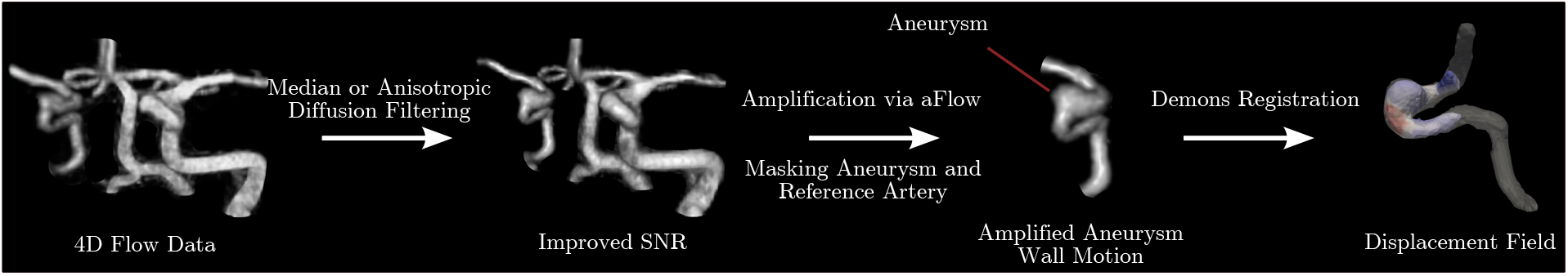
Overview of the data processing and aFlow method: First, 4D flow data were filtered to improve SNR. Then, aneurysms and reference arteries were masked and the 4D flow images were amplified. Finally, the displacement fields were extracted using the Demons registration algorithm.

**Figure 2:**
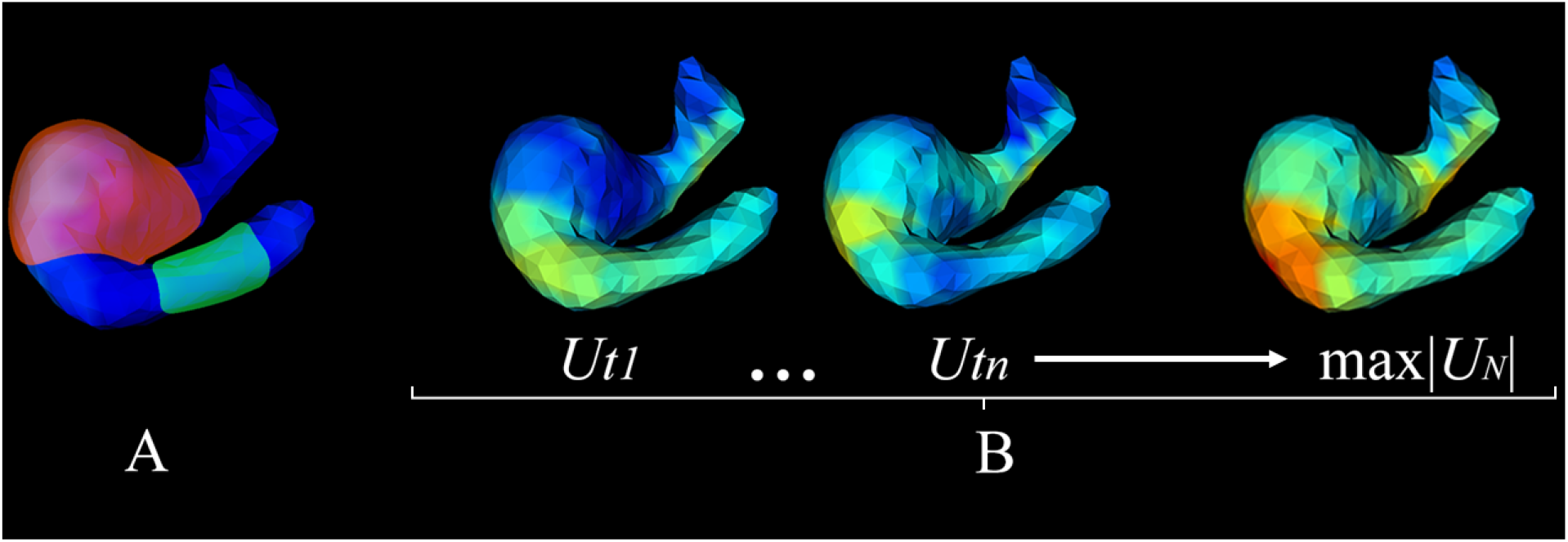
Overview of the displacements measurement. Selection of ROI: aneurysm ROI (red) and healthy artery ROI (green) (A). Extraction of normalized maximum displacement for each time step (*t*_1_ to *t*_*n*_) to generate the volume *t*_*max*_ of maximum normalized displacement over time (B).

**Figure 3:**
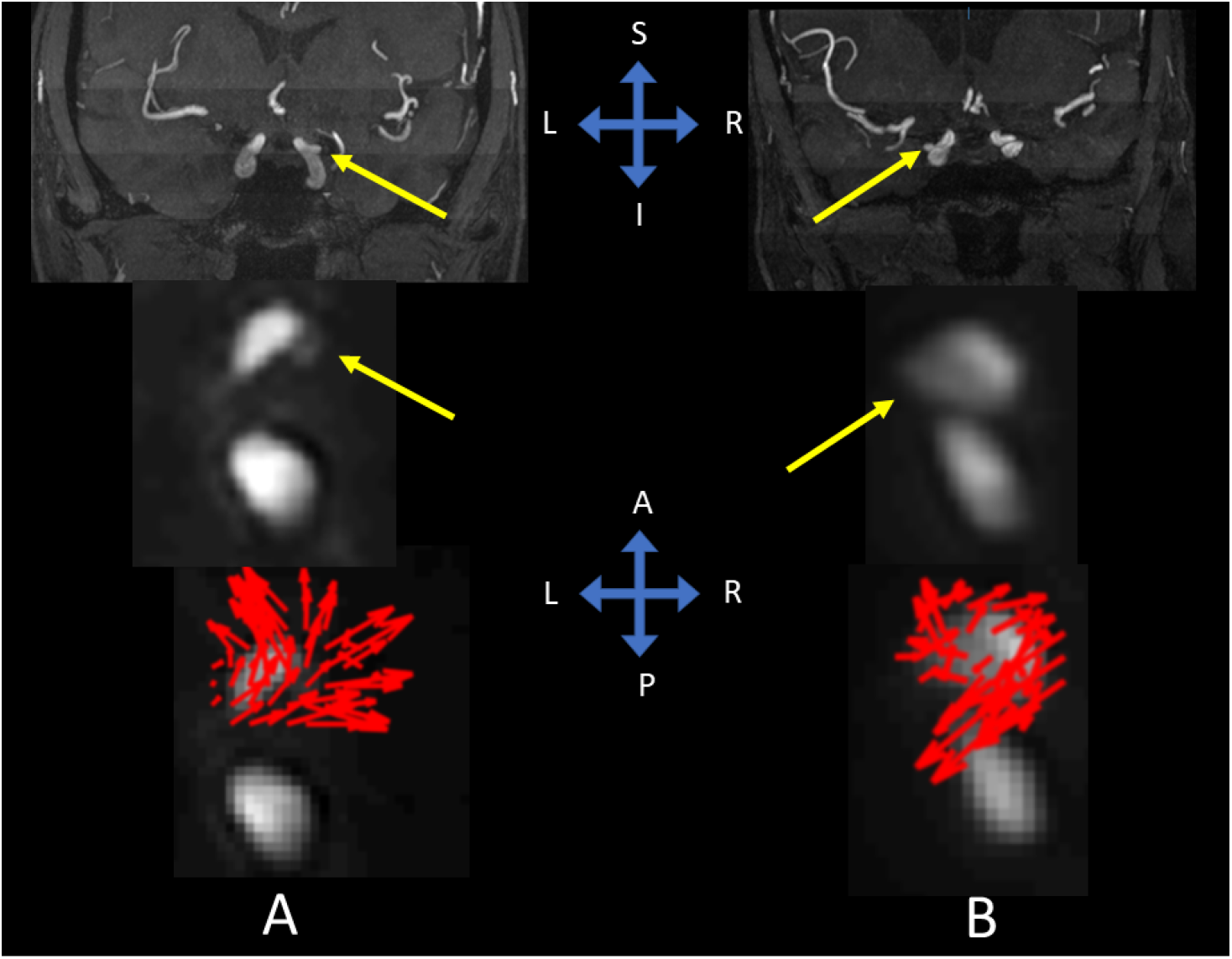
Comparison of the displacement fields between stable (A) and growing aneurysms (B). The first row shows the MRA images of the IAs in the coronal plane. The second row show the 4D flow images of the IAs in the axial plane. The third row shows the displacement fields symbolized by arrows for each IA. The stable aneurysm shows homogeneous displacement field, globally oriented in the same direction and comparable to a uniform bulging of the aneurysm sac. On the contrary, growing aneurysm presents inhomogeneous motions, similar to a swirling motion.

Before applying aFlow on the data, de-noising steps were taken to improve the signal-to-noise ratio (SNR). Here, median [49, 50] and diffusion filtering [51] were performed to improve the contrast between the aneurysm edge and the stationary background. Following this, the Matlab histogram matching algorithm [52] was used between each time step (with respect to the first time step) to reduce the temporal intensity variation, resulting in an improved temporal SNR [53].

### 2.5. aFlow algorithm

To visualize the subtle aneurysm wall displacement in 3 directions, the aFlow algorithm was used on the preprocessed data. In the first step, by using a 3D steerable pyramid, aFlow decomposes the data at each time step into local phases at different scales and orientations [38, 53, 54]. Then, the temporal variations of the decomposed data at each time-step with respect to the first time point was calculated (*i.e*. phase difference at each level and orientation). It should be noted that these phase differences correspond to the local displacement of the preprocessed 4D flow MRI data at each scale and orientation [55]. Next, the DMD modal decomposition technique was used on the local phase differences, allowing one to isolate the motion at specific temporal frequencies and capture the highly transient phenomena present in the aneurysm wall deformation [38, 56]. In short, DMD sums up the modes in the selected range of frequencies making it possible to capture the dominant behavior of a dataset in the absence of its governing model [57–60]. An amplitude-weighted Gaussian spatial filter can be used on the DMD processed phase differences to reduce the noise that might arise from the amplification. The filtered phase data were multiplied by an amplification factor *α* and added back to the original phase data. Finally, the modified 4D flow MRI dataset is reconstructed by collapsing the 3D steerable pyramid, resulting in an amplified dataset in the selected frequency range [38].

### 2.6. Analysis of the in vivo data through aFlow

The aFlow algorithm was applied to the 4D flow MRI datasets in our 20 patients to extract wall motion metrices. To analyze the aneurysm wall motion via the aFlow algorithm, we selected an amplification factor of *α* = 3 and two frequency ranges of *f* ∈ [0 1.5 *f*_*H*_] Hz (denoted as ”main harmonics”) and *f* ∈ [1.5 *f*_*H*_ 3 *f*_*H*_] Hz (denoted as ”higher frequencies”), where *f*_*H*_ corresponds to the heart rate of the analyzed subject.

### 2.7. Displacement measurement

After the amplification, the displacement fields were extracted with the Demons registration algorithm [61, 62]. Then, a region of interest (ROI) containing all the voxels of the aneurysm was selected. Another reference ROI including the parent artery voxels was selected a few mm away from the aneurysm neck. The ROI of the reference artery was used to normalize the displacement of the aneurysm wall for every individual. To calculate the normalized displacement of each aneurysm with respect to the reference artery at the selected ROI, we used the following formulation:

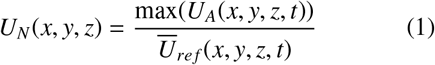

where *U*_*N*_ (*x, y, z*) is the normalized displacement at x, y, z coordinates on the aneurysm wall (the voxels of the aneurysm wall), and max(*U*_*A*_(*x, y, z, t*)) and *Ū*_*ref*_ (*x, y, z, t*) are the maximum aneurysm and mean reference artery displacements over time t, respectively. These voxel-based displacement values were calculated for each aneurysm and used to calculate the following parameters: maximum of *U*_*N*_ (max|*U*_*N*_|), 90^*th*^ percentile of 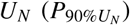, 50^*th*^ percentile of 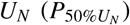, 10^*th*^ percentile of 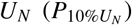, standard deviation of 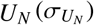, and interquartile range of 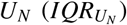. Here, 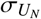 and the 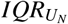 are indicators of the deformation variability across the same aneurysm geometry.

### 2.8. Statistical Analysis

A paired-wise analysis was adopted to assess the differences in clinical variables, demographic data, morphological features, and aFlow parameters between patients with stable versus growing aneurysms. Baseline characteristics and neuroimaging variables were compared between subjects with stable and growing aneurysms employing t-tests, Wilcoxon, or McNemar tests as appropriate. Significant variables following univariate analysis were tested by a conditional (fixed-effect) logistic regression and predictive values were provided as Odds ratios with a 95% confidence interval. Unmatched methods, *e.g*. unconditional logistic regression, were used in addition to our loose-matching data based on the findings of [63]. Finally, the correlation between significant variables was assessed by Pearson correlation analysis. All statistical analyses were carried out at the *p* =0.05 (2-sided) significance level using IBM SPSS Statistics for Windows (Version 24.0) and Stata Statistical Software (Release 15).

### 2.9. CFD Analysis

Time resolved computational fluid dynamic simulation was conducted for 6 stable and 6 growing aneurysms using a similar methodology to that outlined in McGah et al [64]. In this work patient specific velocity inlet boundary conditions were obtained via 4D flow MR velocimetry. The meshes were constructed using unstructured polyhedral elements with a characteristic length of 0.125 mm and six prism layers at all wall surfaces. The average number of elements in each mesh is 1.5 million. Blood was assumed to be an incompressible Newtonian fluid with a density of 1050 kg/m^3^ and a viscosity of 3.5cP. Transient simulations were run for 6 cardiac cycles at a time step of 0.001 seconds to ensure the results are independent of initial conditions. The hemodynamic parameters of interest in this work are the local fluid pressure (P), Wall Shear Stress (WSS), Oscillatory Shear Index (OSI), and Directional Wall Shear Stress Gradient (WSSGdir).

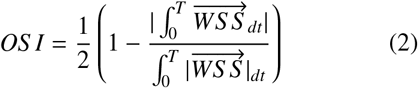

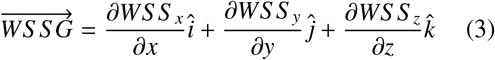

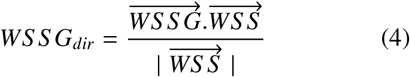

OSI measures the oscillation of WSS by dividing the magnitude of the time average of the WSS vector with the time average of the magnitude of WSS and has been shown to correlate directly with aneurysm instability and rupture [65].

Directional wall shear stress gradient is defined to be positive when the spatial gradient of WSS is in the same direction as the WSS vector, representing an acceleration of flow, while a negative value represents a deceleration. The time variance of WSSGdir therefore is a good measure of the change in direction of flow within the aneurysm, which we hypothesize will correlate with wall motion. In this work we developed 10 representative hemodynamic parameters that correlate with wall motion and aneurysm stability by taking spatial averages, time averages, time coefficient of variance and spatial standard deviations of the pressure, WSS, OSI, and WSSdir. In addition to statistical measures, we examined the percent of the aneurysm dome area that had outlier levels of these variables when compared to the same values calculated for the reference artery. These 10 parameters are defined in Table 1.

**Table 1:**
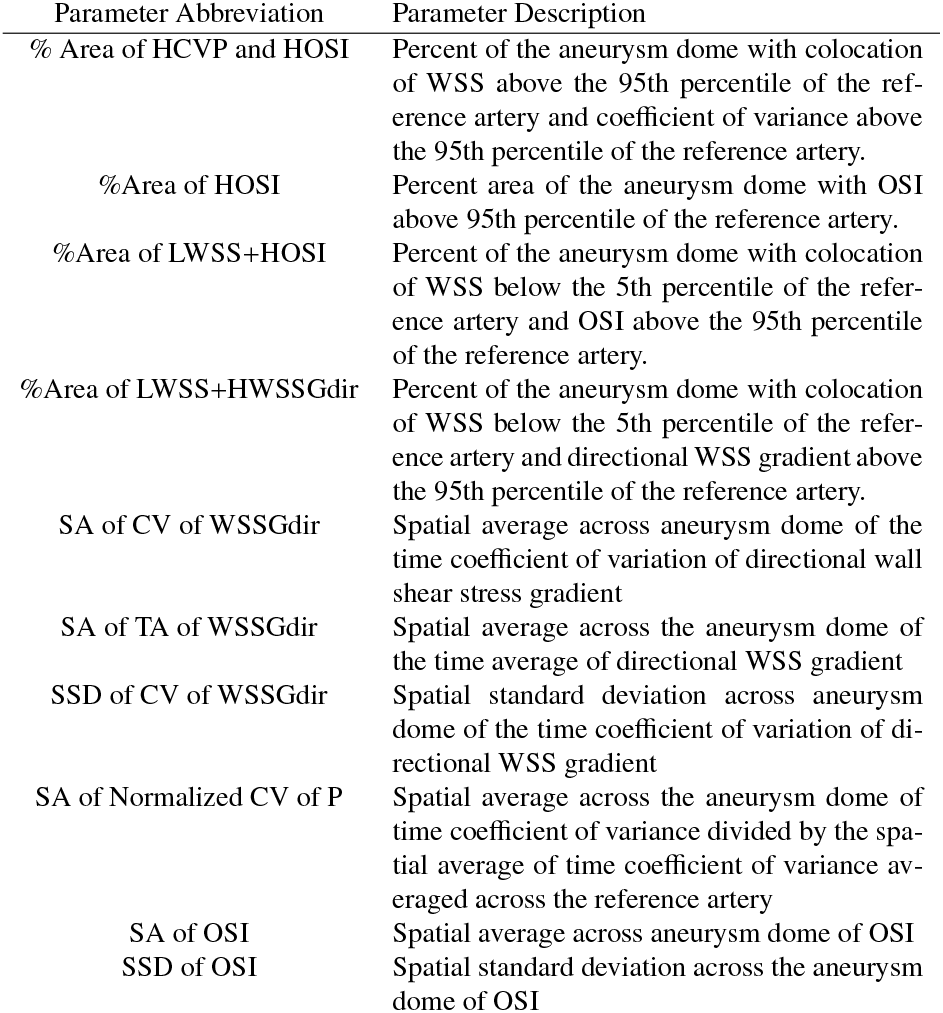
Description of CFD hemodynamic parameters

| Parameter Abbreviation | Parameter Description |
| --- | --- |
| % Area of HCVP and HOSI | Percent of the aneurysm dome with colocation of WSS above the 95th percentile of the reference artery and coefficient of variance above the 95th percentile of the reference artery. |
| %Area of HOSI | Percent area of the aneurysm dome with OSI above 95th percentile of the reference artery. |
| %Area of LWSS+HOSI | Percent of the aneurysm dome with colocation of WSS below the 5th percentile of the reference artery and OSI above the 95th percentile of the reference artery. |
| %Area of LWSS+HWSSGdir | Percent of the aneurysm dome with colocation of WSS below the 5th percentile of the reference artery and directional WSS gradient above the 95th percentile of the reference artery. |
| SA of CV of WSSGdir | Spatial average across aneurysm dome of the time coefficient of variation of directional wall shear stress gradient |
| SA of TA of WSSGdir | Spatial average across the aneurysm dome of the time average of directional WSS gradient |
| SSD of CV of WSSGdir | Spatial standard deviation across aneurysm dome of the time coefficient of variation of directional WSS gradient |
| SA of Normalized CV of P | Spatial average across the aneurysm dome of time coefficient of variance divided by the spatial average of time coefficient of variance averaged across the reference artery |
| SA of OSI | Spatial average across aneurysm dome of OSI |
| SSD of OSI | Spatial standard deviation across the aneurysm dome of OSI |

## 3. Results

### 3.1. Wall motion results

There were no statistical differences among demographic and clinical variables between patients with stable versus growing IAs and no statistical differences in these between the two groups (Table 2). We also compared baseline aneurysm measurements including sac size, neck size, aspect ratio and location (Table 3). Total follow up time (*mean* ± *SD*) was 30.6 ± 9.1 months, 28.4 ± 9 in the stable vs. 32.9 ± 9.1 in the growing cohort. Following the final size measurement analysis, the final aneurysm sac size (*mean* ± *SD*) in the stable cohort was 4.50 ± 2.56 mm (*p* = 0.77, mean difference: 0.03 mm, standard error of mean difference: 0.21 mm). In the growing cohort, the final sac size (*mean* ± *SD*) was increased to 7.53 ± 4.65 mm (*p* = 0.012, mean difference: 1.12 mm, standard error of mean difference: 0.36 mm). A quantitative analysis of IAs wall displacement in stable and growing cohorts are summarized in Table 4.

**Table 2:** Demographic and clinical variables in patients with stable vs. growing aneurysms (univariate analysis).

| Variable | Stable (n=10) | Growing (n=10) | p-value |
| --- | --- | --- | --- |
| Age (years) | 56.5 ± 13.1 | 57 ± 16.9 | 0.16 |
| Sex (M/F) | 1/9 | 3/7 | 0.62 |
| Family history of aneurysm | 4 (40%) | 0 (0%) | 1 |
| Family history of SAH | 2 (20%) | 1 (10%) | 1 |
| History of hypertension | 5 (50%) | 9 (90%) | 1 |
| History of smoking | 3 (30%) | 5 (50%) | 0.12 |
| History of alcohol consumption | 2 (20%) | 3 (30%) | 1 |
| History of migraine | 1 (10%) | 1 (10%) | 1 |
| Aneurysm multiplicity | 4 (40%) | 5 (50%) | 0.37 |
| PHASES score; Median (IQR) | 1 (0.75-2.25) | 1 (0.75-6.25) | 0.3 |

**Table 3:** Baseline aneurysm size and morphometric features. * *p* values are based on paired-wise analysis. ** Data are presented as mean ± SD, unless otherwise specified.

| Variable | Stable (n=10) | Growing (n=10) | p-value** |
| --- | --- | --- | --- |
| Sac size (mm) | 4.48 (2.60) | 5.95 (3.77) | 0.062 |
| Neck size (mm) | 2.86 (1.36) | 3.98 (0.80) | 0.034 |
| Aspect Ratio | 1.71 (0.91) | 1.46 (0.81) | 0.328 |
| Location (ICA/MCA) (number) | 8/2 | 8/2 | 1.00 |

**Table 4:**
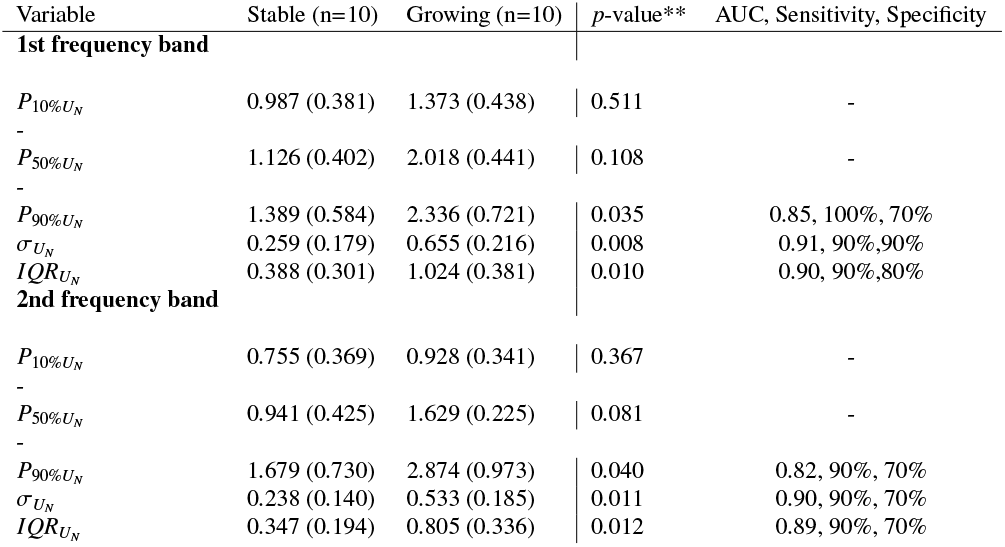
aFlow parameters in patients with stable vs. growing aneurysms, first and second frequency bands. * *p* values are based on paired-wise analysis. ** Data are presented as mean ± SD, unless otherwise specified.

| Variable | Stable (n=10) | Growing (n=10) | p-value** | AUC, Sensitivity, Specificity |
| --- | --- | --- | --- | --- |
| **1st frequency band** |  |  |  |  |
| $P_{10\%U_N}$ | 0.987 (0.381) | 1.373 (0.438) | 0.511 | - |
| - | - | - | - | - |
| $P_{50\%U_N}$ | 1.126 (0.402) | 2.018 (0.441) | 0.108 | - |
| - | - | - | - | - |
| $P_{90\%U_N}$ | 1.389 (0.584) | 2.336 (0.721) | 0.035 | 0.85, 100%, 70% |
| $\sigma_{U_N}$ | 0.259 (0.179) | 0.655 (0.216) | 0.008 | 0.91, 90%, 90% |
| $IQR_{U_N}$ | 0.388 (0.301) | 1.024 (0.381) | 0.010 | 0.90, 90%, 80% |
| **2nd frequency band** |  |  |  |  |
| $P_{10\%U_N}$ | 0.755 (0.369) | 0.928 (0.341) | 0.367 | - |
| - | - | - | - | - |
| $P_{50\%U_N}$ | 0.941 (0.425) | 1.629 (0.225) | 0.081 | - |
| - | - | - | - | - |
| $P_{90\%U_N}$ | 1.679 (0.730) | 2.874 (0.973) | 0.040 | 0.82, 90%, 70% |
| $\sigma_{U_N}$ | 0.238 (0.140) | 0.533 (0.185) | 0.011 | 0.90, 90%, 70% |
| $IQR_{U_N}$ | 0.347 (0.194) | 0.805 (0.336) | 0.012 | 0.89, 90%, 70% |

Using the acquired displacement fields, we calculated 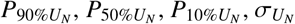, and 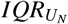 for each subject at two frequency ranges. Overall, we observed significantly higher 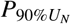 values and wider 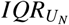 and 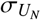 values in growing IAs as compared to the stable group regardless of the frequency (*p* < 0.05, Table 4). We found that on average, 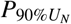 in both of the frequency bands are approximately 70% higher in the growing IAs (Figure 4(C), Table 4). The maximum wall IAs motions depicted by 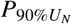 values were significantly higher in the growing IAs in comparison to the stable IAs (*p* = 0.035 for the 1^*st*^ frequency and 0.04 for the 2^*n*^ frequency wall motion).

**Figure 4:**
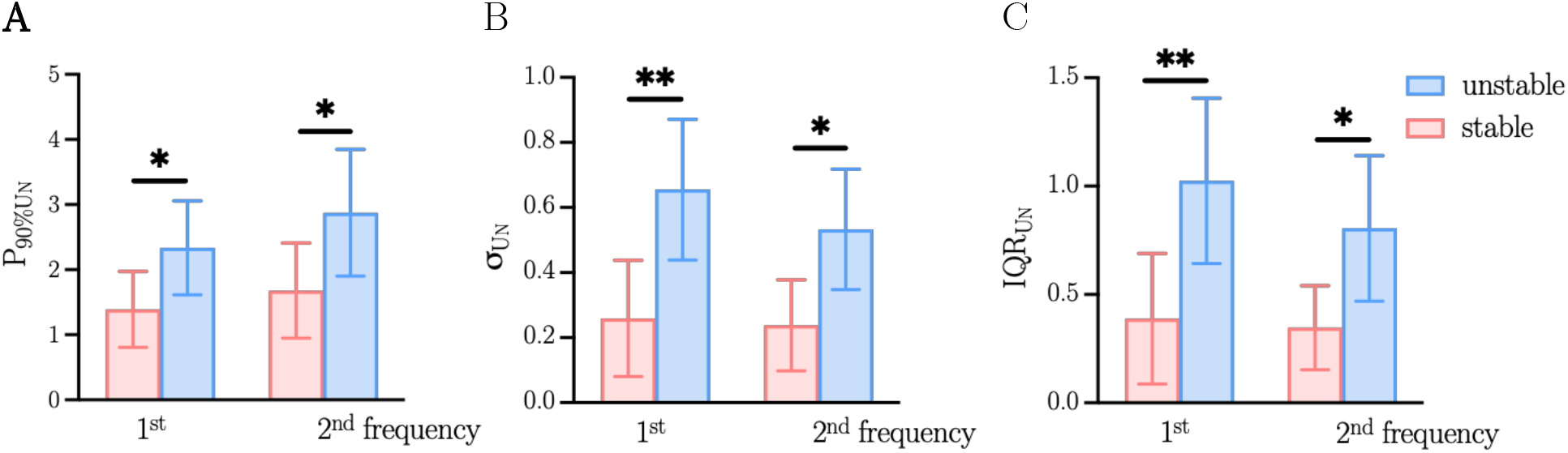
Comparison of displacement parameters between stable and growing aneurysms. 90^*th*^ percentile of normalized displacement 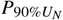 (A), standard deviation 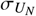 (B) and interquartile range 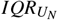 (C)

The growing IAs also showed higher variability of deformation across their geometry, evident by the dispersion variables including 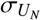 and 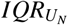. We found approximately 153% and 124% larger 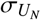 in the 1^*st*^, and the 2^*nd*^ frequency ranges of the growing IAs as compared to the stable ones, respectively (Table 4). These values for 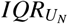 were 164% and 132% larger in the 1^*st*^ and the 2^*nd*^ frequency ranges of the growing IAs, respectively (Fig. 4.D, Table 4). There was no significant difference in the IAs wall motion when considering 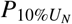 or 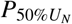between stable vs. growing IAs (Table 4). Similar results were observed for the 2nd frequency band. The same parameters were significantly higher for growing IAs: 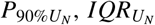 and 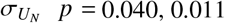 and 0.012, respectively).

### 3.2. CFD Results

After analyzing the hemodynamic results, we found several hemodynamic parameters correlate with wall motion, and are themselves predictive of aneurysm stability (Figure 5). In order to reduce transient, 3-dimensional data into single parameters we examined time averages (TA), spatial averages (SA), time coefficients of variance (CV) and spatial standard deviations (SSD) of hemodynamic parameters. We also examined the percent area of the aneurysm dome that experienced outlier levels of OSI, variance in pressure, WSS and WSSGdir. An outlier level was defined as greater than the 95th percentile (or lower than 5th percentile) of the value of the same parameter in the reference artery. Table 5 shows the correlation between 10 hemodynamic parameters and measures of wall motion and aneurysm stability. The best predictor of maximum wall motion and variance of wall motion is the spatial standard deviation of the time coefficient of variance of the directional wall shear stress gradient. This backs up our hypothesis that aneurysms with large changes in the direction of WSS will also have higher levels of wall motion. The percent of the dome that has a colocation of outlier high levels of OSI and outlier high levels of time coefficient of variation of pressure is also highly correlated with wall motion. The areas with high oscillation in WSS, but with low levels of WSS will be those with vorticity and rotational flow, and as such it makes sense they would correlate with wall motion. Table 5 also compares the hemodynamic parameters for the stable and growing aneurysms. The best hemodynamic discriminator from a Receiver Operator Characteristic (ROC) perspective is the spatial standard deviation of the coefficient of variance of directional wall shear stress gradient, matching the finding from the wall motion analysis that spatial variance is the best predictor of aneurysm growth. The spatial average of the time average of *WSSG*_*dir*_ is also a strong predictor of aneurysm growth, with high values corresponding to growing aneurysms. These high values of *WSSG*_*dir*_ corresponds with flow accelerating into the aneurysm dome, which intuitively should correlate with aneurysm growth. While the directional wall shear stress gradient variables perform well in an ROC analysis, the t-test reveals that the difference in means between stable and unstable aneurysms is not quite statistically significant. Neither the spatial average or spatial variance of OSI was a significant predictor of aneurysm growth, but the percent area of the aneurysm with outlier high levels of OSI is a statistically significant predictor. This indicates that, similar to wall motion, it is important to consider OSI relative to the base state of the reference artery when examining the impact on aneurysm growth. The spatial average across the aneurysm dome of the coefficient of variance of pressure, normalized by the spatial average of the same value across the reference artery, was a statistically significant predictor of growing aneurysms. A high value of normalized coefficient of variance of pressure indicates that the pressure is fluctuating more in the dome of the aneurysm than the rest of the artery on average, and this increase in pressure fluctuation correlates well with growing aneurysms. Furthermore, the most statistically significant difference in means was in the percent of aneurysm dome area with a colocation of outlier high OSI and high variance in pressure. This indicates that normal and shearing stress both fluctuating at high rates in the same region leads to aneurysm growth. Figure 6 displays contours of the areas of colocation of elevated variance in pressure and elevated OSI for the stable and unstable cohorts.

**Table 5:**
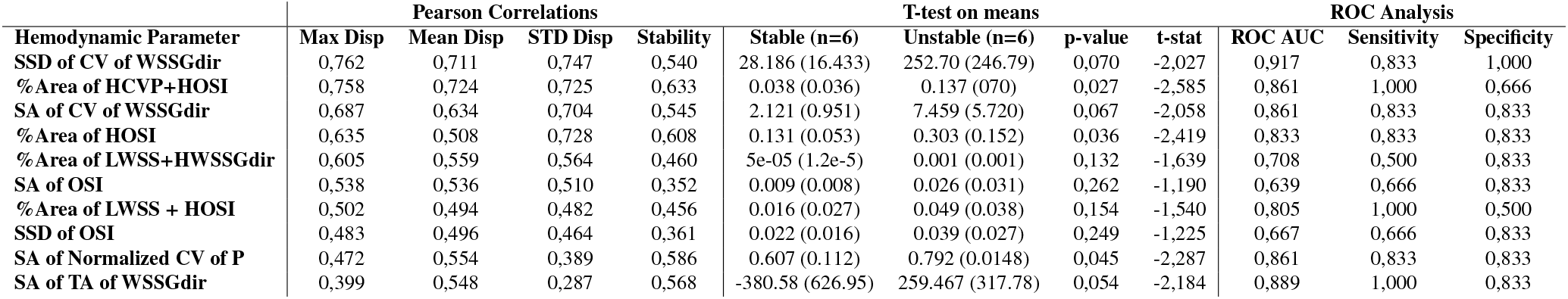
Pearson correlations between hemodynamic parameters and wall motion, and t-test and ROC statistics between stable and unstable aneurysms for the hemodynamic parameters.

| Hemodynamic Parameter | Pearson Correlations |  |  |  | T-test on means |  |  |  | ROC Analysis |  |  |
| --- | --- | --- | --- | --- | --- | --- | --- | --- | --- | --- | --- |
|  | Max Disp | Mean Disp | STD Disp | Stability | Stable (n=6) | Unstable (n=6) | p-value | t-stat | ROC AUC | Sensitivity | Specificity |
| SSD of CV of WSSGdir | 0,762 | 0,711 | 0,747 | 0,540 | 28.186 (16.433) | 252.70 (246.79) | 0,070 | -2,027 | 0,917 | 0,833 | 1,000 |
| % Area of HCVp+HOSI | 0,758 | 0,724 | 0,725 | 0,633 | 0.038 (0.036) | 0.137 (070) | 0,027 | -2,585 | 0,861 | 1,000 | 0,666 |
| SA of CV of WSSGdir | 0,687 | 0,634 | 0,704 | 0,545 | 2.121 (0.951) | 7.459 (5.720) | 0,067 | -2,058 | 0,861 | 0,833 | 0,833 |
| % Area of HOSI | 0,635 | 0,508 | 0,728 | 0,608 | 0.131 (0.053) | 0.303 (0.152) | 0,036 | -2,419 | 0,833 | 0,833 | 0,833 |
| % Area of LWSS+HWSSGdir | 0,605 | 0,559 | 0,564 | 0,460 | 5e-05 (1.2e-5) | 0.001 (0.001) | 0,132 | -1,639 | 0,708 | 0,500 | 0,833 |
| SA of OSI | 0,538 | 0,536 | 0,510 | 0,352 | 0.009 (0.008) | 0.026 (0.031) | 0,262 | -1,190 | 0,639 | 0,666 | 0,833 |
| % Area of LWSS + HOSI | 0,502 | 0,494 | 0,482 | 0,456 | 0.016 (0.027) | 0.049 (0.038) | 0,154 | -1,540 | 0,805 | 1,000 | 0,500 |
| SSD of OSI | 0,483 | 0,496 | 0,464 | 0,361 | 0.022 (0.016) | 0.039 (0.027) | 0,249 | -1,225 | 0,667 | 0,666 | 0,833 |
| SA of Normalized CV of P | 0,472 | 0,554 | 0,389 | 0,586 | 0.607 (0.112) | 0.792 (0.0148) | 0,045 | -2,287 | 0,861 | 0,833 | 0,833 |
| SA of TA of WSSGdir | 0,399 | 0,548 | 0,287 | 0,568 | -380.58 (626.95) | 259.467 (317.78) | 0,054 | -2,184 | 0,889 | 1,000 | 0,833 |

**Figure 5:**
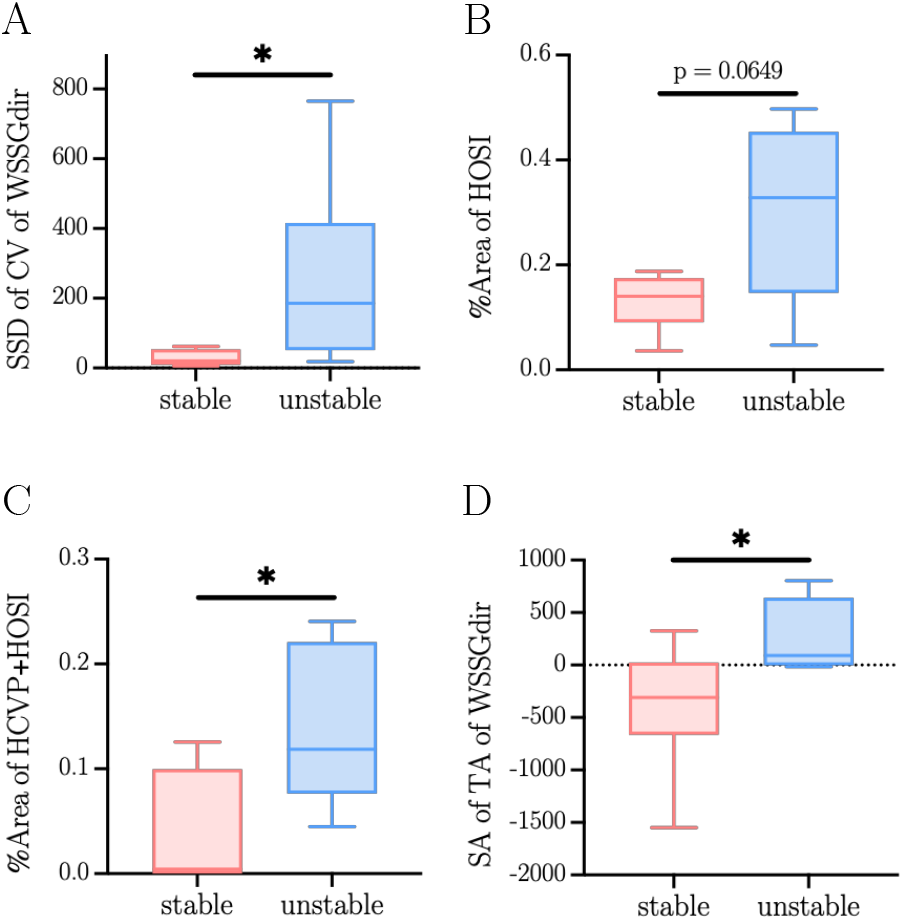
Comparison of hemodynamic parameters between stable and growing aneurysms. SSD of CV of WSSGdir (A), %Area of HOSI (B), %Area of HCVP+HOSI (C) and SA of TA of WSSGdir (D)

**Figure 6:**
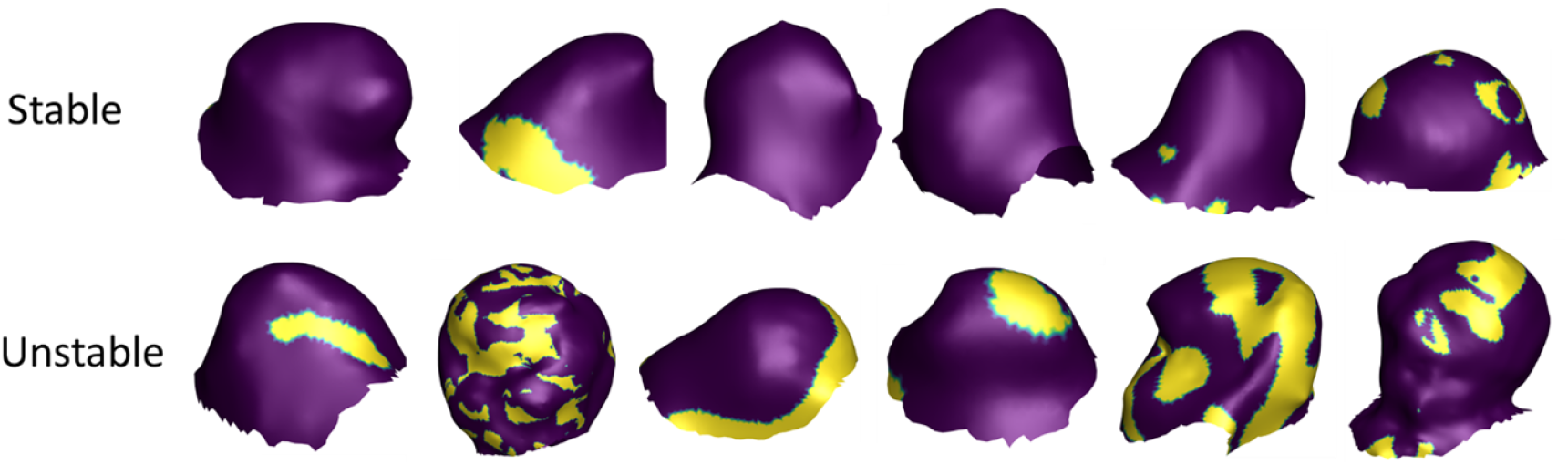
Colocation of elevated OSI and elevated coefficient of variation of pressure. Contour plots of colocation of elevated OSI and elevated coefficient of variation of pressure for stable and unstable aneurysms. Colocation areas are shown in yellow, while area without colocation are shown in purple.

## 4. Discussion

This work showed that aFlow can provide quantitative analysis of IAs wall motion and differentiate growing IAs from stable IAs at baseline imaging with high diagnostic accuracy in a pairwise analysis. We specifically showed that growing IAs have inherently higher wall motion and more heterogenous wall deformation compared to stable IAs.

A heterogenous pattern of motion and wall deformation has been associated with growing and ruptured IAs [29, 30]. Conventional *in vivo* imaging techniques were previously used to quantify aneurysm wall motion [30, 34–37]. However, such small deformations in wall motion are close to the imaging resolution, making these deformations difficult to visualize, thereby rendering these approaches unreliable in the clinical setting [66]. Currently, four imaging techniques are often used to measure aneurysm wall motion: power Doppler ultrasonography, phase-contrast MRA, 3D rotational catheter angiography and 4D-CTA [30] each with some technical and practical limitations. Observed pulsation with transcranial ultrasonography is operator dependent and depends on the device setting, which raises concerns about reproducibility [67]. Both 3D rotational catheter angiography and 4D-CTA can achieve high spatial and temporal resolutions although with practical limitations for a non-invasive screening tool related to requirements for radiation and injection of contrast material [30]. Phase-contrast MRA is an appealing alternative due to lack of radiation and no requirements for contrast injection, however it can suffer from low spatial resolution and flow artifacts from out-of-range velocities [50]. aFlow, however, allows one to clearly visualize the cerebrovasculature wall motion during the cardiac cycle, while also achieving sufficient spatial and temporal resolution for wall motion quantification [38].

In this study, our recent results using aFlow showed that the maximum IAs wall motion depicted by 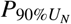 values were significantly higher in the growing IAs in comparison to the stable IAs. This is concordant to prior reports suggesting that growing and ruptured aneurysms were subject to higher pulsatile motion [31, 32]. Neurosurgical assessment confirmed that the IAs rupture points are the sites of pulsating blebs [31] and also match the location of the observed wall motion abnormality during preoperative analysis [32]. High pulsatile motion of the wall of intracranial aneurysms could be due to several factors, such as its complex geometry and its interactions with the blood flow [68].

Indeed, abnormal flow patterns such as turbulent flows are associated with high risk aneurysms [56, 69, 70]. Moreover, flow related metrics such as oscillatory shear index or wall shear stress obtained via computational fluid dynamics have been correlated with risks of rupture and growth [71]. These flow related abnormalities may result in high motion of the aneurysm wall. Therefore it is plausible that capturing high arterial wall motion could be an indicator of the presence of underlying flow related abnormalities, and that useful diagnostic metrics of this motion can be obtained through aFlow without the need for complicated CFD analysis.

Another important finding in our results is that 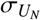 and 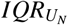 dispersion measures were also significantly higher in growing aneurysms in comparison to the stable aneurysms (Table 4), suggesting that growing IAs have higher heterogeneity of wall motion with areas of high and low motion resulting in wide dispersion metrics. The causes of these dyskinetic movements may have multiple origins, such as specific blood flow patterns or inhomogneous mechanical properties of the aneurysm wall, previously proven in the literature to be related to the risk of rupture. The association between unusual hemodynamics, increased risk of rupture [72, 73] and blebs formation [74, 75] has been shown. These abnormal flow patterns are characterized by large and concentrated inflow jets, complex and oscillatory flow patterns, and wall shear stress distributions with focal regions of high shear and large regions of low shear stress, which likely result in inhomogeneous aneurysm wall motion. From the perspective of tissue mechanical characterization, analysis of wall thickness and other parameters such as yield stress and strain in aortic aneurysms showed that different regions of the aneurysm have different mechanical properties [76]. In cerebral aneurysms, rupture seems to be caused by localized degradation and weakening of the wall [77]. Indentation tests on an intracranial aneurysms sample also found that the ruptures occured in a restricted area of increased elastic capacity and that unruptured areas had increased stiffness [78]. While to our knowledge no direct causal link has been demonstrated between variation in mechanical properties and IAs wall deformation, it can be reasonably assumed that regions of lower stiffness are subject to higher deformation. These localized areas of higher or lower wall stiffness are likely causing the inhomogeneous wall motion. The complementary CFD analyses performed on a subset of stable and growing aneurysms were in line with our assumptions. The correlation between hemodynamic parameters and wall motion parameters seemed to confirm our hypothesis that large changes in WSS direction would lead to large deformations. Furthermore, the best hemodynamic discriminator of stability was spatial variance, which supported the observations made in this study and in prior reports that growing aneurysms have more heterogeneous wall motions. Finally, the results showed that wall motion parameters are equivalent or better predictors than hemodynamic parameters for the assessment of aneurysm stability, which was a very encouraging result that allows to consider aFlow as a potential substitute for CFD analyses, although the number of samples is insufficient to draw significant conclusions. Our study has several limitations. The retrospective nature introduces unknown bias. Despite our best efforts to collect prospective data for nearly 5 years, the sample size remains small. This is an inherent limitation of any study of this type where the required outcome (*i.e*. aneurysm growth) has relatively low incidence. Moreover, the majority of growing aneurysms were likely to get treated and were not enrolled in this type of wait and watch practice. Unlike our previous study [38], no significant differences were observed between the two frequency bands in determining instability. In the current study, displacement metrices from both frequency bands were significantly correlated with growth (Table 4). This is probably due to the smaller number of imaging phases in the current dataset. We anticipate that increasing the number of cardiac phases and improvement in temporal resolution can probably unmask the potential added value of higher harmonics. The differences between information obtained from low and high frequency bands require further investigation.

Although we showed that other clinical and demographic data were not contributing to growth prediction, this should be interpreted in the context of selection bias in matching the control cohort. After all, patients in the control cohort were matched based on aneurysm size and location, both of which have been considered significant risk factors for IAs. Another limitation is related to potential differences in aneurysm wall motion depending on location and geometry of the aneurysms. We did however normalize the values based on parent artery for each aneurysm to calculate normalized values and mitigate heterogeneity. Also the analysis was pairwise, *i.e*. MCA aneurysm displacement metrices were compared against another MCA aneurysm in the control group to mitigate differences due to IAs locations.

Finally, the aFlow algorithm can suffer from technical limitations related to noise and artifacts. Motion artifacts can occur if a high amplification factor is used or if the unamplified motion is initially too large [38, 53]. However, amplification parameters were calibrated with phantoms and *in vivo* testing [38] to mitigate this limitation.

## 5. Conclusion

In this study, we used the aFlow algorithm to amplify the subtle motions of the cerebrovasculature, to test the ability of aFlow to differentiate stable from unstable IAs. The wall motion of IAs were assessed in patients with growing aneurysms and compared to those with stable aneurysms. A CFD analysis was also performed on a subset of stable and growing aneurysms. Higher aneurysm wall motion and higher variability of deformation was shown in the growing aneurysms compared to the stable aneurysms.These differences are possibly due to inhomogeneities of the mechanical characteristics of the vessel walls or to underlying hemodynamics, which were correlated with aneurysms instability and risk of rupture in previous studies. CFD analysis results confirmed these observations.

We have demonstrated here that the measurement and amplification of the aneurysm wall motion achieved with aFlow has the potential to differentiate stable from growing aneurysms. These results are encouraging and, with further validation, we believe that this technique could be a valuable tool for the assessment of aneurysm stability and potentially act as a substitute for in depth computational fluid dynamic analysis.

## Data Availability

All data produced in the present study are available upon reasonable request to the authors

## Notes

### Competing Interest Statement

The authors have declared no competing interest.

### Funding Statement

This study did not receive any funding

### Author Declarations

IRB of Icahn School of Medicine at Mount Sinai gave ethical approval for this work

